# Quantifying delay in first contact with HIV programs among young women engaged in sex work in Mombasa, Kenya: a time-to-event analysis

**DOI:** 10.1101/2025.03.29.25324768

**Authors:** Huiting Ma, Kristy CY Yiu, Linwei Wang, Griffins Manguro, Peter Gichangi, Helgar Musyoki, Parinita Bhattacharjee, Shem Kaosa, Japheth Kioko, Shajy Isac, Chidumebi Idemili, James Blanchard, Marissa Becker, Sharmistha Mishra

## Abstract

**Background:** Young women engaged in sex work (YSW) experience a disproportionately high burden of HIV, yet most HIV programs for sex workers are not designed to reach adolescent girls and young women.

**Methods:** Longitudinal data from the start of sex work are infeasible, but cross-sectional surveys may help identify “contact gaps”. We used data from the 2015 *Transitions* Study, a cross-sectional survey of sexually active women aged 14-24 who self-identified as a sex worker in Mombasa, Kenya. We created a virtual cohort using self-reported event timing described relative to the survey date. We quantified the time from self-identification as sex worker to the initial program contact (“contact gap”) and employed time-to-event analyses to estimate and characterize factors associated with the rate of program contact.

**Results:** Of 392 YSW, 47 (12%) reported program contact, with a median time of 12 months (interquartile range: 2, 24). The rate of program contact per 100 person-months was 0.52 [95% confidence interval (CI): 0.38, 0.68], and when applied to the estimated population size of YSW in Mombasa, the minimum access gap was 11,532 person-years. A shorter contact gap was associated with: older age when first negotiated for sex (adjusted hazard ratio:1.2 [95%CI: 1.0, 1.5]); and self-perceived ease of earning money through sex work (11.5 [2.8, 47.7]).

**Conclusion:** A large contact gap highlights the need to reshape HIV prevention services for YSW across their life course. Despite limitations, cross-sectional data could help estimate the contact gap and support program monitoring and evaluation.

## INTRODUCTION

Women engaged in sex work experience a disproportionately high burden of HIV (1), with 13.5-fold HIV prevalence compared to other women of reproductive age (1, 2). Data suggest that young women engaged in sex work (YSW, aged 14 to 24 years) may be more vulnerable to HIV acquisition than their older counterparts (3), with more than two-thirds of HIV acquisition occurring before or during the first few years after self-identification as a sex worker (4). For example, in South Africa the HIV incidence among YSW aged 18-24 was almost three times of sex workers aged 25 and above (5). In Nairobi, Kenya, 12% of women had been diagnosed with HIV within the first two years of sex work (6).

Data are emerging on HIV risks and gaps in reach of HIV prevention programs at the intersections of sex work and adolescence (3, 7–11). Studies suggest that YSW report larger number of clients and lower levels of condom use as compared to older women in sex work (11–16). Reaching YSW with HIV prevention services shortly after they first enter into ‘formal’ sex work and/or self-identify as sex workers has been challenging for programs and for YSW (17, 18) (11, 15). First, HIV prevention programs designed to reach a broader community of adolescent girls and young women may not address the specific needs of YSW in the context of vulnerabilities related to sex work (7–11). Second, most HIV prevention programs tailored for women in sex work were historically designed under legal parameters to provide services for women over the age of 18 who self-identify as sex workers (11, 15, 19, 20). Taken together, YSW have had limited access to a range of healthcare services and have been described to be “largely invisible” in HIV prevention programmatic initiatives and research (11, 21, 22).

To adequately estimate person-time the gaps in the program “reach” in the context of sex work, and longitudinal data from the start of sex work would be needed. Such data are not available, nor would they be feasible to collect in the real world. However, what are available are cross-sectional data that programs traditionally use to measure program “reach”. In the context of HIV programs tailored for women in sex work, “reach” is usually defined as the proportion of sex workers contacted by peer educators (23–25) and provides an overall snapshot of program contact for the estimated population of women in sex work, at a specific time point. Given the the increasing awareness of vulnerabilities experienced prior to program contact (26), programs would also benefit from understanding the magnitude of person-time of vulnerabilities and associations between vulnerabilities and program contact.

Due to the lack of longitudinal data, we sought to leverage cross-sectional survey data among YSW, which included questions on the timing of specific events, to help address the above knowledge gap. Specifically, we used cross-sectional data among YSW aged 14-24 recruited via sex work venues (hotspots) in Mombasa, Kenya in 2015, to generate a pseudo cohort using self-reported event timing described relative to the survey date. Our specific objectives were to: i) quantify the time from self-identifying as a sex worker to initial contact with a program tailored for women in sex work (contact gap); and ii) identify determinants of program contact conceptualized along the timeline of sex work using time-to-event analyses.

## METHODS

### Study setting and population

Data were obtained from the *Transitions* Study, a cross-sectional biological and behavioral survey conducted between April and November 2015 in Mombasa, Kenya,, among: cis-gender women aged 14-24 years frequenting sex work venues (hotspots) (27); who had ever engaged in vaginal or anal sex; and provided written informed consent. In 2014, the HIV prevalence in Mombasa was estimated at 6.0% and 10.0% among females in the age groups 15-19 and 20-24, respectively (26). There were estimated to be 6,127 (range 4,793-7,462) women engaged in sex work between the ages of 14 and 24 years in Mombasa in 2014, representing approximately 4.5% of the overall female population of the same age group (27).

We restricted the current analyses to participants who self-identified as a sex worker at the time of the survey. We discerned participants as self-identified sex workers by asking the following questions in the survey: “Presently, do you consider yourself a sex worker?” or “Were you ever a sex worker?”

### Data collection

Details of the community mobilization, survey protocol, and data collection were previously reported by Cheuk et al. (27, 28). Briefly, participants were recruited using probability proportional to the estimated population size of YSW at the hotspots (27, 28) those who were eligible and consented, and those who were eligible and consented received face-to-face surveys with a structured survey in English or Kiswahili (**Appendix 1**). We transformed cross-sectional data into a pseudo cohort using self-reported timing described in reference to the survey date (**Appendices 2 and 3**).

### Measures

Programs were defined as services provided by local non-governmental organizations, community-based organizations, and/or faith-based organizations. We examined two levels of self-reported program engagement: (a) program contact as our primary outcome; and (b) program access as our secondary outcome. We defined program contact as the outreach activity of peer educators when they first meet with sex workers. Peer educators are current or former sex workers who work with the local HIV prevention program for FSW to provide outreach to sex workers at hotspots. We defined program access as self-reported use of any of the following services offered by FSW programs: condom use education and demonstrations; provision of clean needles/syringes; program clinic or drop-in center services such as testing for sexually transmitted infections or HIV.

We defined the contact gap as the time from self-identification as a sex worker to initial program contact. For the secondary outcome, we defined the access gap as the time from self-identification as a sex worker to program access.

We conceptualized covariates related to contact and access gaps along the timeline of engagement in sex work (**Appendix 1**) and existing literature (7, 29–33). Covariates related to age at specific events of interest were identified and the remaining were categorized into two time periods; the first time period includes covariates that occurred from the time up to, at, and during the first month of sex work, and the second time period includes the current perceptions and lifetime covariates. **Figure 1** depicts the conceptual framework and **Table 1** and **Appendix 4** lists the covariates.

**Figure 1.**
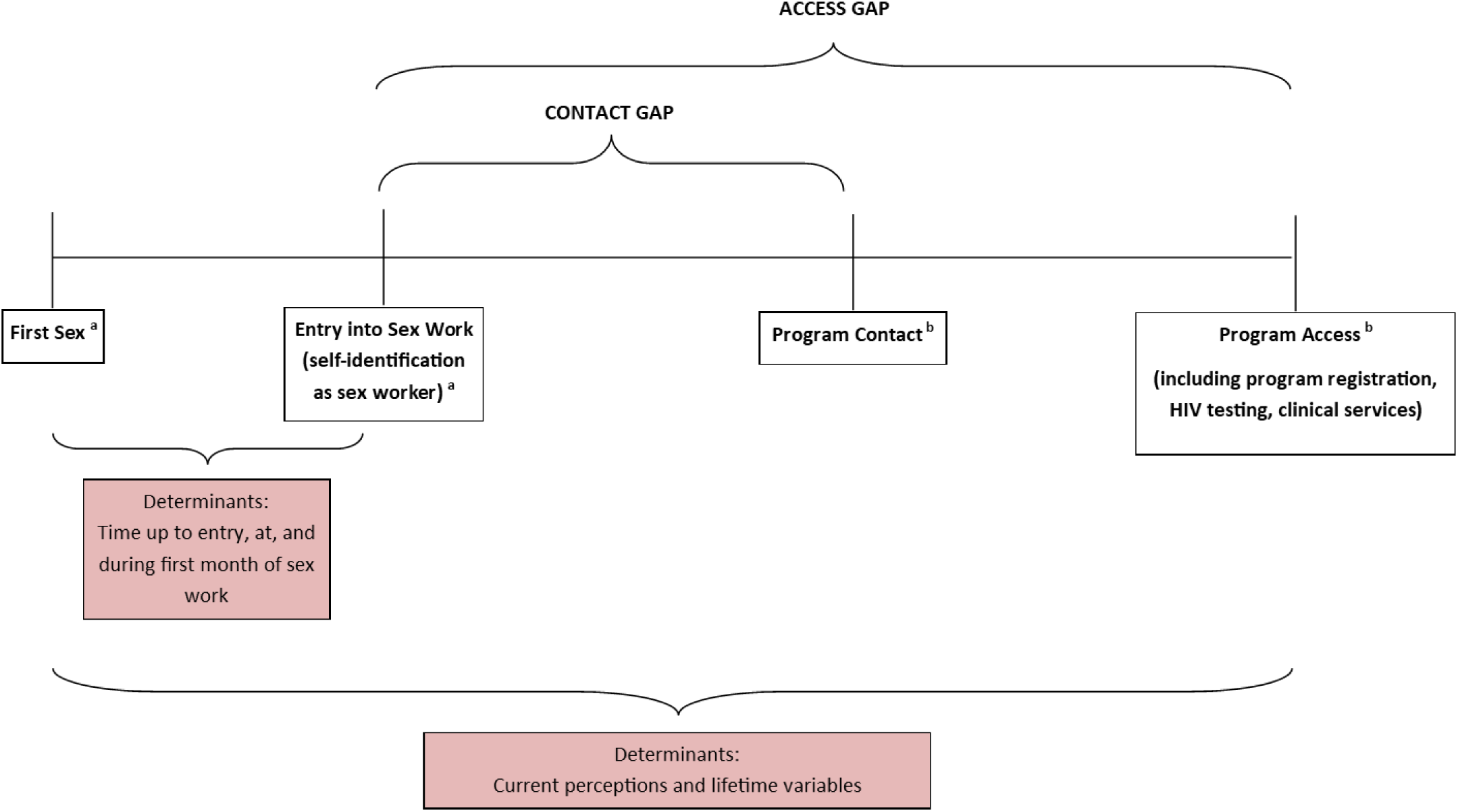
Conceptual framework. The diagram illustrates the sequential order of events described in the study. ^a^ The events of first sex and self-identification as a sex worker may not occur in sequential order. It is possible that the two events occurred concurrently.^b^ Participants who were contacted by programs or have accessed programs prior to self-identification as sex workers were excluded from time-to-event analyses.

**Table 1.** Characteristics of study participants between those who have ever/never been contacted by program by time of study

| Characteristics (N(%)) | Total (N = 408) | Contacted by program by time of study (N = 61) | Never contacted by program (N = 347) | P-value |
| --- | --- | --- | --- | --- |
| **AGE AT EVENT OF INTEREST** |  |  |  |  |
| Current age, years (Median [IQR]) | 20 (18-22) | 20 (19-22) | 20 (18-22) | 0.07 |
| Age at self-identification as sex worker, years (Median [IQR]) | 18 (16-20) | 18 (17-20) | 18 (16-20) | 0.65 |
| Age at first negotiation for sex, years (Median [IQR]) | 18 (16-19) | 18 (17-19.5) | 18 (16-19) | 0.10 |
| **TIME UP TO ENTRY, AT, AND DURING FIRST MONTH OF SEX WORK** |  |  |  |  |
| First sex before age 15 | 138 (33.8%) | 21 (34.4%) | 117 (33.7%) | 1.00 |
| Exchanged money at first sex | 129 (31.6%) | 12 (19.7%) | 117 (33.7%) | 0.04 |
| Forced sex at first sex | 59 (14.5%) | 6 (9.8%) | 53 (15.3%) | 0.33 |
| Tested for HIV by entry into sex work | 212 (52.0%) | 36 (59.0%) | 176 (50.7%) | 0.27 |
| History of pregnancy by entry into sex work | 131 (32.1%) | 16 (26.2%) | 115 (33.1%) | 0.27 |
| Aware of program by entry into sex work | 32 (7.8%) | 19 (31.1%) | 13 (3.7%) | <0.001 |
| Experienced police violence any time up to entry, at, and during first month of sex work | 60 (14.7%) | 9 (14.8%) | 51 (14.7%) | 1.00 |
| Experienced physical violence any time up to entry, at, and during first month of sex work | 37 (9.1%) | 3 (4.9%) | 34 (9.8%) | 0.33 |
| Experienced forced sex any time up to entry, at, and during first month of sex work | 71 (17.4%) | 4 (6.6%) | 67 (19.3%) | 0.02 |
| Monthly living expenses covered by first month of sex work | 43 (10.5%) | 4 (6.6%) | 39 (11.2%) | 0.37 |
| **CURRENT PERCEPTIONS AND LIFETIME VARIABLES** |  |  |  |  |
| Self-perceived difficulty of earning money through sex work<sup>i</sup> |  |  |  | <0.001 |
| Difficult (1-3) | 123 (30.1%) | 8 (13.1%) | 115 (33.1%) |  |
| Neutral (4-6) | 98 (24%) | 11 (18%) | 87 (25.1%) |  |
| Easy (7-10) | 180 (44.1%) | 41 (67.2%) | 139 (40.1%) |  |
| Sex worker identity known, ever | 384 (94.1%) | 58 (95.1%) | 326 (93.9%) | 1.00 |
| Told others sex worker identity, ever | 294 (72.1%) | 43 (70.5%) | 251 (72.3%) | 0.76 |
| Tested for HIV, ever | 382 (93.6%) | 59 (96.7%) | 323 (93.1%) | 0.40 |
| Self-perceived to be at moderate/great risk of HIV infection | 222 (54.4%) | 33 (54.1%) | 189 (54.5%) | 1.00 |
| Regularity in current source of income | 67 (16.4%) | 9 (14.8%) | 58 (16.7%) | 0.85 |

### Statistical analyses

We compared the characteristics of study participants with and without program contact using Chi-squared or Fisher’s exact tests for categorical covariates when appropriate and the Kuskal-Wallis test for continuous covariates. We used descriptive statistics to quantify the length of the contact gap. We further examined the characteristics of experiencing other vulnerabilities prior to entry to sex work and used descriptive statistics to quantify the time from experiencing those vulnerabilities to self-identification as sex workers and to program contact, respectively.

We quantified the rate of program contact and assessed the relationships between program contact and covariates in the following time-to-event analyses. Individuals who were contacted by programs prior to self-identification as sex workers (N=12) and those who did not report time from self-identification as sex workers to program contact (N=4) were excluded in time-to-event analyses (**Appendix 5**). We used the Kaplan-Meier estimator to estimate the survival function of program contact for: 1) all study participants overall; and 2) study participants stratified by covariates of interest. The log rank test was used to compare survival functions by covariates and two-sided *P* values were calculated. We also calculated the rate of contact (per 100 person-months) for each survival function.

Cox proportional hazard regression models (34) with and without adjustment for age at the time of survey (i.e., described as current age in the survey) were applied to estimate hazard ratios (HRs) and the 95% confidence intervals (CIs)(35). We used the participant’s age at the time of the survey in the age-adjusted Cox regression because the covariates examined were closely associated with our outcomes and age.

We repeated our descriptive analyses and time-to-event analyses for our secondary outcome (program access).

Finally, we triangulated the minimum number of person-years of program contact delay among all YSW in Mombasa, assuming our study sample is representative of the YSW in Mombasa, using the following: estimated number of YSW in Mombasa (27) and the estimated number of person-months of delays from our study sample (details in **Appendix 6**).

R version 4.0.2 was used for all analyses and graphics (36). *P* values <0.05 were considered to be statistically significant.

### Ethics approval

The Human Research Ethics Board at the University of Manitoba, Canada (HS16557 [H2013:295]); and the Kenyatta National Hospital-University of Nairobi Ethical Review Committee, Kenya (P497/10/2013) provided institutional ethics approval. Research permit was received from the National Commission for Science, Technology and Innovation, Kenya.

## RESULTS

### Participant characteristics (Table 1)

Of the 408 participants who met the inclusion criteria, the median age at the time of the survey was 20 (interquartile range [IQR]: 18, 22). The median age at first self-identification as sex worker was 18 (IQR: 16, 20). A total of 15% (61/408) of participants were contacted by programs at the time of survey.

A total of 129 (31.6%) participants reported that their first sex included exchange of money for sex. Participants with program contact were less likely to have exchanged money at first sex (33.7% vs. 19.7%, *P*=0.04). Participants with program contact were less likely to have experienced forced sex by first month of sex work (6.6% vs. 19.3%, *P*=0.03). There was no statistically significant difference between participants with and without program contact with respect to experience of physical violence by first month of sex work. Of 59 who experienced forced sex by first months of sw, first experience of forced sex occurred a median of 12 months (IQR: 1.5, 24) prior to sex work. Only 6/59 (10.2%) contacted by program, with a median of 8 months from first experience of forced sex to program contact.

Overall, 384 participants reported that friends and relatives were aware of participants’ engagement in sex work, and 294 participants (72.1%) had disclosed and told others they self-identified as a sex worker. A total of 180 (44.1%) viewed earning money through sex work as easy. Participants with program contact were more likely than those without program contact to perceive earning money through sex work as easy (67.2% vs. 40.1%, *P*<0.001).

### Patterns of program contact

Of the 408 participants, 392 (96.1%) answered the question related to time to program contact during the study period and did not report contacted by program before sex work were included in the time-to-event analysis. Forty-seven (12%) reported program contact, of whom, the median contact time was 12 months (interquartile range:2, 24) (**Appendix 5A**). The rate of program contact per 100 person-months was 0.52 (95% confidence interval: 0.38, 0.68) (**Figure 2A**). Compared to those who did not exchange money at first sex, those who exchanged money had a lower rate of program contact (0.31 vs. 0.62 per 100 person-months; log rank test *P*=0.05) and a longer median length of contact gap (19 months vs. 17 months) (**Figure 2B**). Compared to those who perceived earning money through sex work as difficult, those who perceived earning money through sex work as easy had a higher rate of program contact (0.86 vs. 0.07 per 100 person-months; log rank test *P*<0.001) and a longer median length of contact gap (19 months vs. 17 months) (**Appendix 7**).

**Figure 2.**
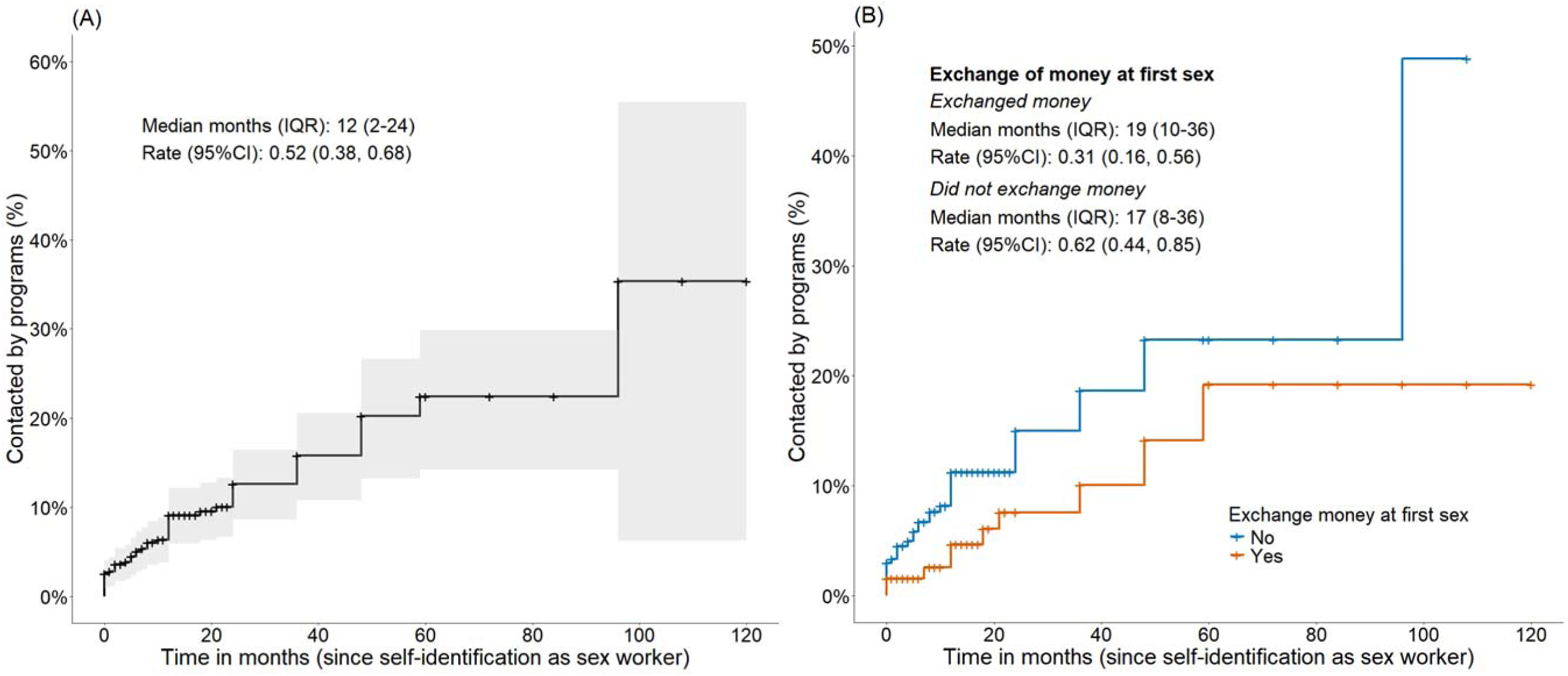
Kaplan-Meier curves of time from self-identification as sex worker to program contact for all participants (A) and exchange of money at first sex. The graphs depict length of contact gap (time in months from initial self-identification as a sex worker to initial program contact). **Abbreviations:** CI: confidence interval; IQR: interquartile range

### Determinants of program contact (Figure 3)

After adjusting for current age, older age when first negotiated money in exchange for sex (adjusted hazard ratio [aHR]: 1.2; 95% CI: 1.0, 1.5; *P=*0.021), not exchanged money at first sex (aHR: 0.5; 95% CI: 0.3, 1.1; *P=*0.082), aware of program by entry into sex work (aHR: 6.4; 95% CI: 2.9, 14.2; *P*<0.001), and self-perceived difficulty of earning money through sex work as easy (aHR: 11.5; 95% CI: 2.8, 47.7; *P<*0.001) and neutral (aHR: 5.1; 95% CI: 1.1, 23.9; *P=*0.041) vs. as difficult were associated with increased hazards of program contact, indicating a shorter contact gap.

**Figure 3.**
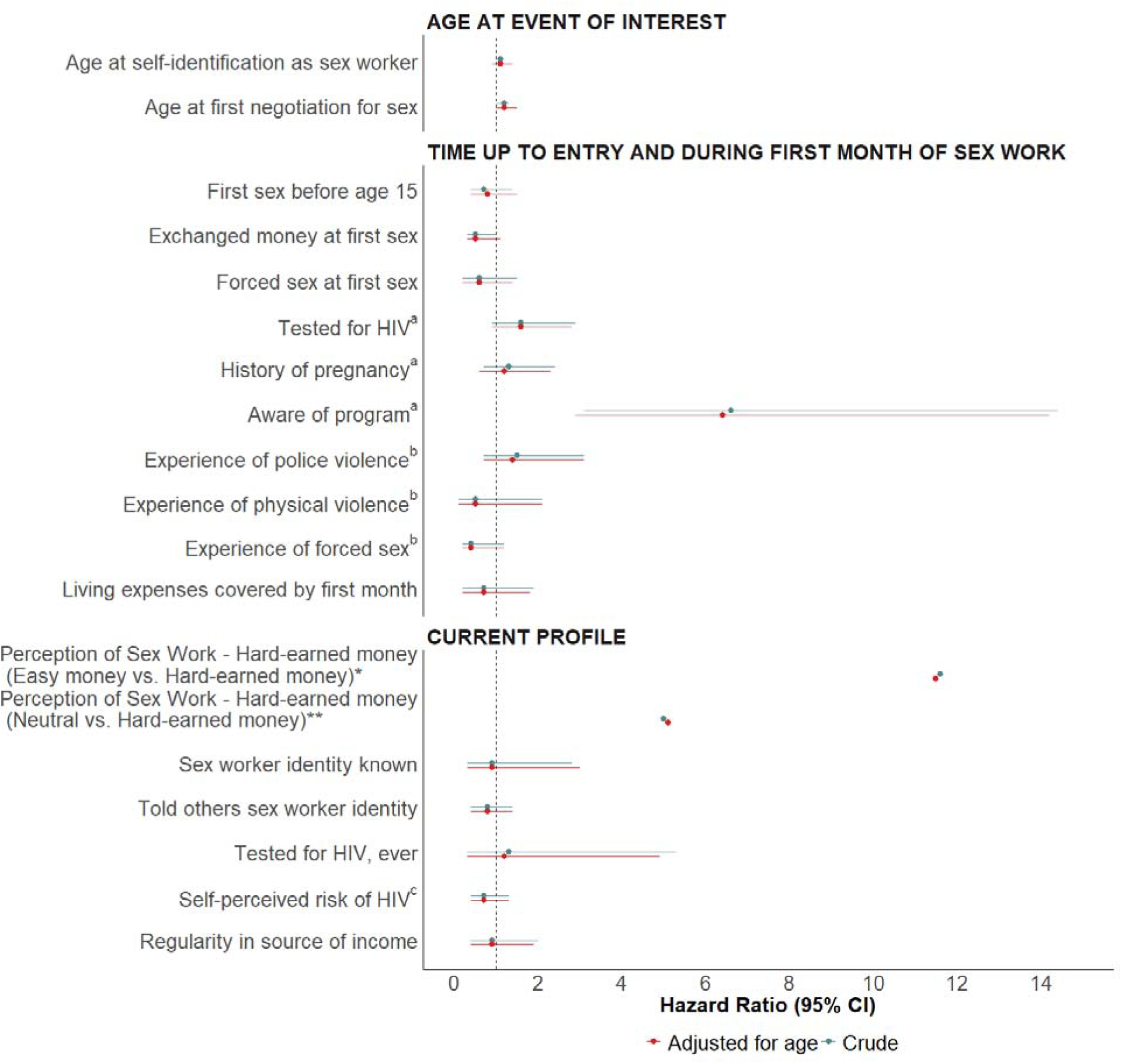
Forest plot of non-adjusted and adjusted hazard ratios by determinants on contact gap. Figure depicting association between determinants and time to program contact. Hazard ratios are adjusted for current age. **Abbreviation:** CI: confidence interval ^a^ By entry into sex work. ^b^ Any time up to entry, at, and during first month of sex work. ^c^Self-perceived risk of HIV is stratified into participants who perceived their risk of HIV to be moderate/great and those who perceived the risk to be small/none/unsure (reference).*Crude hazard ratio: 11.6; 95% CI: 2.8, 48.3; adjusted hazard ratio: 5.0; 95% CI: 1.1, 23.7. **Crude hazard ratio: 5.1; 95% CI: 1.1, 23.9; adjusted hazard ratio: 5.1; 95% CI: 1.1, 23.9).

### Population-level contact gap for all YSW in Mombasa

Based on the estimate of 6,127 YSW between the ages of 14 and 24 years in Mombasa in 2014 (28), we estimated a minimum of 11,532 person-years contact gap among all FSW in Mombasa (Appendix 6).

### Patterns of program access (Table 2, Appendices 8-10)

A total of 56 participants had accessed program services. Participants with program access were less likely to have experienced forced sex (5.4% vs. 19.3%, *P*=0.008) by the first month of sex work, and more likely to aware of progam by entry into sex work (33.9% vs. 3.7%, *P*<0.001). Among the 53 participants who reported time from self-identification as FSW to program access, the rate of program access per 100 person-months was 5.7 (95% CI: 4.3, 7.4), and the median length to program access among those who had access was 8 months (IQR: 1, 24). After adjusting for age, the determinants are similar to the determinants of contact gap, and the following were associated with a shorter access gap: self-perceived difficulty of earning money through sex work as easy vs. difficult (aHR: 3.6; 95%CI: 1.5, 8.6; *P*=0.004), did not experience sexual violence by first month of sex work (aHR: 0.3; 95%CI: 0.1, 0.9; *P*=0.037), and aware of program by entry into sex work (aHR: 13.9; 95%CI: 7.6, 25.5; *P*<0.001). Exchange of money at first sex was slightly associated with a longer access gap (aHR: 0.5; 95%CI: 0.2, 1.0; *P*=0.04).

**Table 2.** Characteristics of study participants between those who have ever/never accessed the program by time of study

| Characteristics (N(%)) | Total (N = 408) | Accessed program (N = 56) | Never accessed program (N = 352) | P-value |
| --- | --- | --- | --- | --- |
| **AGE AT EVENT OF INTEREST** |  |  |  |  |
| Current age, years (Median [IQR]) | 20 (18-22) | 20 (19-22) | 20 (18-22) | 0.07 |
| Age at self-identification as sex worker, years (Median [IQR]) | 18 (16-20) | 18 (17-20) | 18 (16-20) | 0.94 |
| Age at first negotiation for sex, years (Median [IQR]) | 18 (16-19) | 18 (17-19.5) | 18 (16-19) | 0.18 |
| **TIME UP TO ENTRY, AT, AND DURING FIRST MONTH OF SEX WORK** |  |  |  |  |
| First sex before age 15 | 138 (33.8%) | 19 (33.9%) | 119 (33.8%) | 1.00 |
| Exchanged money at first sex | 129 (31.6%) | 11 (19.6%) | 118 (33.5%) | 0.04 |
| Forced sex at first sex | 59 (14.5%) | 5 (8.9%) | 54 (15.3%) | 0.30 |
| Tested for HIV by entry into sex work | 212 (52.0%) | 32 (57.1%) | 180 (51.1%) | 0.47 |
| History of pregnancy by entry into sex work | 131 (32.1%) | 14 (25.0%) | 117 (33.2%) | 0.28 |
| Aware of program by entry into sex work | 32 (7.8%) | 19 (33.9%) | 13 (3.7%) | < 0.001 |
| Experienced police violence any time up to entry, at, and during first month of sex work | 60 (14.7%) | 8 (14.3%) | 52 (14.8%) | 1.00 |
| Experienced physical violence any time up to entry, at, and during first month of sex work | 37 (9.1%) | 1 (1.8%) | 36 (10.2%) | 0.04 |
| Experienced forced sex any time up to entry, at, and during first month of sex work | 71 (17.4%) | 3 (5.4%) | 68 (19.3%) | 0.008 |
| Monthly living expenses covered by first month of sex work | 43 (10.5%) | 4 (7.1%) | 39 (11.1%) | 0.49 |
| **CURRENT PERCEPTIONS AND LIFETIME VARIABLES** |  |  |  |  |
| Self-perceived difficulty of earning money through sex work |  |  |  | < 0.001 |
| Difficult (1-3) | 123 (30.7%) | 6 (10.9%) | 117 (33.8%) |  |
| Neutral (4-6) | 98 (24.4%) | 11 (20%) | 87 (25.1%) |  |
| Easy (7-10) | 180 (44.9%) | 38 (69.1%) | 142 (41%) |  |
| Sex worker identity known, ever | 384 (94.1%) | 53 (94.6%) | 331 (94%) | 1.00 |
| Told others sex worker identity, ever | 294 (72.1%) | 17 (30.4%) | 255 (72.4%) | 0.64 |
| Tested for HIV, ever | 382 (93.6%) | 54 (96.4%) | 328 (93.2%) | 0.56 |
| Self-perceived to be at moderate/great risk of HIV infection<sup>i</sup> | 222 (54.4%) | 31 (55.4%) | 191 (54.3%) | 1.00 |
| Regularity in current source of income | 67 (16.4%) | 8 (14.3%) | 59 (16.8%) | 0.85 |

## DISCUSSION

We found that 15% of participants reported program contact with a median length of 12 months from self-identifying as a sex worker to program contact, reflecting a rate of 0.52 per 100-person months. After extrapolating this to the entire YSW population in Mombasa, the minimum population estimate of the contact gap was 11,532 person-years. After adjusting for current age, our findings demonstrated that older age when first negotiated money in exchange for sex, awareness of programs by the time of entry into sex work, and perceptions that money earned through sex work as easy vs. difficult was associated with a shorter contact gap; while the exchange of money at first sex was associated with a longer contact gap.

Our finding demonstrate a large gap in the provision of HIV prevention services along the sexual and occupational life-course of women in sex work. This is despite data from national programs that indicated HIV prevention programs tailored for sex workers were achieving >80% levels of reach among women over age 18 who were engaged in sex work (37). By estimating the person-years of the contact gap, our findings signal critical opportunities for the timing of prevention and program reach; and for integration of services that tailor to the unmet needs at the intersection of sex work and adolescence and youth.

Indeed, in 2018, Kenya officially designed policies to include YSW in programs tailored for sex workers (38). Over the last decade, programs across settings have been developed and designed for YSW and young key populations (39).

Our approach and findings suggest that the following approaches could help in the routine monitoring and evaluation of programs for YSW. First, one indicator to collect in routine program data is the contact gap, by asking about when individuals first self-identified as sex workers workers and recording the time of first program contact. Second, as cross-sectional, behavioral, and bio-behavioral surveys are a mainstay in HIV surveillance in the context of key populations(41, 42), our findings suggest it is feasible and valuable to incorporate a minimal set of questions to calculate the contact gap. In the current context of wide-scale disruptions and dismantling of programs for key populations across the global south (40), including data collection, optimizing surveillance efforts with a minimal set of questions to identify “who is left behind and when” offers an actionable strategy to narrow in on person-time of gaps in place of coverage.

Factors associated with the longer contact gap in our study reinforce findings from other studies that have identified heightened risks of HIV and HIV-related vulnerabilities among sex workers who reported earlier age of entry into sex work and who reported transactional first sex. Across contexts, early age of entry into sex work has been associated with increased risk of HIV infection (41, 42), police arrests (41), experience of physical and sexual violence and verbal abuse from paying partners (8), substance use (43), precarious housing and food insecurity (44). Transactional first sex has been associated with forced and/or coerced first sex, economic and food insecurity, structural violence stemming from gender-based power differential, and future experience of future gender-based violence (7, 45, 46).

We found that self-perceptions surrounding earning money through sex work as easy (vs. difficult), and awareness of programes was associated a shorter contact gap. The former may reflect perceptions of sex work as pathways for occupational and financial independence, and thus, potentially less stigmatizing and thus leading to fewer barriers to first contact with peer educators (47) (48). We also found that YSW with program contact had less negative extreme perceptions of sex work when compared to those not yet contacted by programs, suggesting that negative perceptions toward sex work may be a possible barrier for program contact. It may also indicate that through program contact, YSW have developed a more positive perception of sex work as they receive more support. Finally, our findings suggest that increasing general awareness of programs tailored for women in sex work remains an important early component alongside the availability of services.

Our study focused on contact with programs tailored for women in sex work. YSW have other means of accessing HIV services through other componets of the wider HIV prevention programming. These include facility-based HIV testing programs in government facilities, antencare visitsatalvisits, or general outpatient health care visits (49). Indeed, we found that many YSW reported prior HIV testing despite absence of contact with programs tailored for sex workers. This finding demonstrates that, as with many health services in Keny and across the global south, different pillars within the healthcare system serve the needs of communities in different ways. Many women in sex work, not just YSW, obtain HIV testing through government facilities as wl as sex work focused programs (50). At times, such “vertical” programming can fill gaps but also lead to gaps, and one of the key challenges and opportunities with the integration of services across population-specific needs will be to minimize gaps.

Limitations of our study include the inherent limitation of converting cross-sectional data to generate longitudinal estimates. Thus, the length of the contact gap in person-years among the YSW population in Mombasa was likely underestimated as individuals who had not been contacted by time of survey were right censored and their contact delay was underestimated using time from self-identification as sex worker to time of survey. Given the nature of the study (converting cross-sectional to a virtual cohort), individuals’ right censoring may not be independent of study characteristics as those with shorter sex work duration were more likely to be censored given the study design. Second, the time between two events (e.g., time from self-identifying as a sex worker and program contact) was determined based on participants’ recall of past events and the time of each event relative to each other, which are prone to recall bias and could influence the results in either direction. Finally, there was potential for social desirability bias as the data was collected via in-person surveys; as our expectation would a bias towards overstating program contact, it means our findings may be an underestimate of the person-years of contact gap.

In Mombasa, Kenya, in 2015, there was an estimated 11,532 person-years of a contact gap between the time women in sex work self-identify as a sex worker and are reached by programs tailored for sex workers. Despite their limitations, cross-sectional data could help estimate this critical gap, using a life-course perspective, and guide the monitoring and evaluation of programs for YSW.

## Data Availability

All data produced in the present study are available upon reasonable request to the authors

## Author affiliations

Department of Public Health and Primary Care, Faculty of Medicine and Health Sciences, Ghent University, Belgium (Peter Gichangi); Division of Infectious Diseases, Department of Medicine, University of Toronto, Toronto, Canada (Sharmistha Mishra); India Health Action Trust, New Delhi, India (Shajy Isac); International Centre for Reproductive Health, Kenya (Griffins Manguro, Peter Gichangi); Institute for Global Public Health, Department of Community Health Sciences, Rady Faculty of Health Sciences, University of Manitoba, Winnipeg, Canada (Parinita Bhattacharjee, Shajy Isac, James Blanchard, Marissa Becker); MAP Centre for Urban Health Solutions, St. Michael’s Hospital, Unity Health Toronto, Toronto, Canada (Kristy CY Yiu, Huiting Ma, Linwei Wang, Chidumebi Idemili, Sharmistha Mishra); Ministry of Health, National AIDS & STI Control Program (NASCOP), Nairobi, Kenya (Helgar Musyoki); Partners for Health and Development in Africa, Nairobi, Kenya (Shem Kaosa, Japheth Kioko); Technical University of Mombasa, Mombasa, Kenya (Peter Gichangi); Institute of Health Policy, Management and Evaluation, Dalla Lana School of Public Health, University of Toronto, Toronto, Canada (Chidumebi Idemili).

## Acknowledgments

We thank the study participants, field staff, peers and outreach workers, and surveyors. We thank the International Centre for Reproductive Health (Mombasa, Kenya), the National AIDS Control Council and the National AIDS & STI Control Programme (Kenya), the Institute for Economics and Forecasting National Academy of Sciences of Ukraine, the Ukrainian Institute for Social Research after Oleksandr Yaremenko (Kyiv, Ukraine) and their respective staff for support in data collection, dissemination, and knowledge translation activities. We thank the Kenya Technical Support Unit for data-entry and data-cleaning support. We thank Dr. Shajy Isac (University of Manitoba) who led the mapping and enumeration, and Dr. Eve Cheuk (University of Manitoba) who led overall coordination of data collection. We thank all members of the *Transitions* Study Team.

Conflict of interest None declared

**Appendix**: Quantifying delay in first contact with HIV programs among young women engaged in sex work in Mombasa, Kenya: a time-to-event analysis

## Appendix 1. Questions from *Transitions* Study surveys used in variable creation

This study consists of two time-to-event analyses: time from self-identification as sex worker to program contact; and time from self-identification as sex worker to program access. This appendix describes how the following three variables were created: self-identification as sex worker; program contact; and program access.

The variables can be created from multiple questions that aimed to obtain the same information from the participant but were phrased differently. This allows us to circumvent the problem of missing data in variable creation through data imputation. The missing data is substituted with the response of a similar question.

Variables: Self-identification as sex worker

- How many years ago did you first consider yourself a sex worker?
  - If 2 years ago or less, in what month and year did you first consider yourself a sex worker?
- How many years ago did you first enter sex work?
  - If 2 years ago or less, in what month and year did you first enter sex work?
- For how long have/had you been in sex work?
- For how long have you been taking paying clients?
- How many years ago was it when you first had sex with a man where the price of sex was negotiated before the sex event?
  - If 2 years ago or less, in what month and year did you first have sex with a man where the price of sex was negotiated before the sex event?

Variables: Program contact

- Are you aware of any NGOs, CBOs or FBOs working on the prevention of HIV/AIDS for young women AND/OR female sex workers in
- How old were you when you first knew about this (these) NGO(s), CBO(s) or FBO(s)?
- How many years ago were you first contacted by peers/staff from an NGO, CBO or FBO?
  - If 2 years ago or less, in what month and year were you first contacted by peers/staff from an NGO, CBO or FBO?
- Approximately how long after you had your first paying client were you first contacted by peers/staff from an NGO, CBO OR FBO?
- Approximately how long after you first considered yourself a sex worker were you first contacted by peers/staff from an NGO, CBO OR FBO?

Variable: Program access

- Are you registered with an NGO, CBO, and FBO?
- When was the FIRST TIME you saw a demonstration on correct use of a male condom by a peer educator, outreach worker or staff from an NGO, CBO or FBO?
- When was the FIRST TIME you were given clean needles/syringes by a peer/worker from an NGO, CBO or FBO?
- Have you ever used any clinics run by an NGO, CBO or FBO?
- Approximately how long after you had your first paying client did you first use any clinics run by an NGO, CBO or FBO?
- Approximately how long after you first considered yourself a sex worker did you first use any clinics run by an NGO, CBO or FBO?
- How many times have you used these clinics to see the doctor in the LAST MONTH?
- When was the FIRST TIME you used the services in any drop-in centres run by an NGO, CBO or FBO?
- Are you a peer worker/peer educator of an NGO, FBO or CBO?

## Appendix 2. Outcome definitions

### A.2.1 *Outcome event: ever contacted by the program by* the time of interview (ever_contact)

<u>Main question</u> (please see **Appendix 1** for detailed questions and possible answers)

- Q803: How many years (and months) ago were you first contacted by peers/staff from an NGO, CBO or FBO?
  a. ever_contact=1, if they reported a valid time since they were contacted by a program (N=56);
  b. ever_contact=0, if they reported never been contacted by peers/staff from a program (N=344);
  c. ever_contact=1, if participants reported they don’t know/don’t remember the time since they were contacted by a program (N=4);
  d. ever_contact=missing, otherwise (N=4).

Among 311 ever_contact=missing, we used the following questions to reduce missing values:

- Q804: Approximately how long after you had your first paying client were you first contacted by peers/staff from an NGO, CBO OR FBO?
- Q805: Approximately how long after you first considered yourself a sex worker were you first contacted by peers/staff from an NGO, CBO OR FBO?
  a. ever_contact=missing => ever_contact=1(N=1), if participants reported a valid time between first paying client and first contacted by peers/staff from a program OR time since the participant first self-identification as sex worker to first program contact ever_contact=missing=> ever_contact=0(N=3), if participants did not provide any of the answers above

**Final:** N=61/408 participants reported that they have ever been contacted by a program by the time of interview

### A.2.2 Time-to-event (Time from self-identification as sex worker to first program contact)

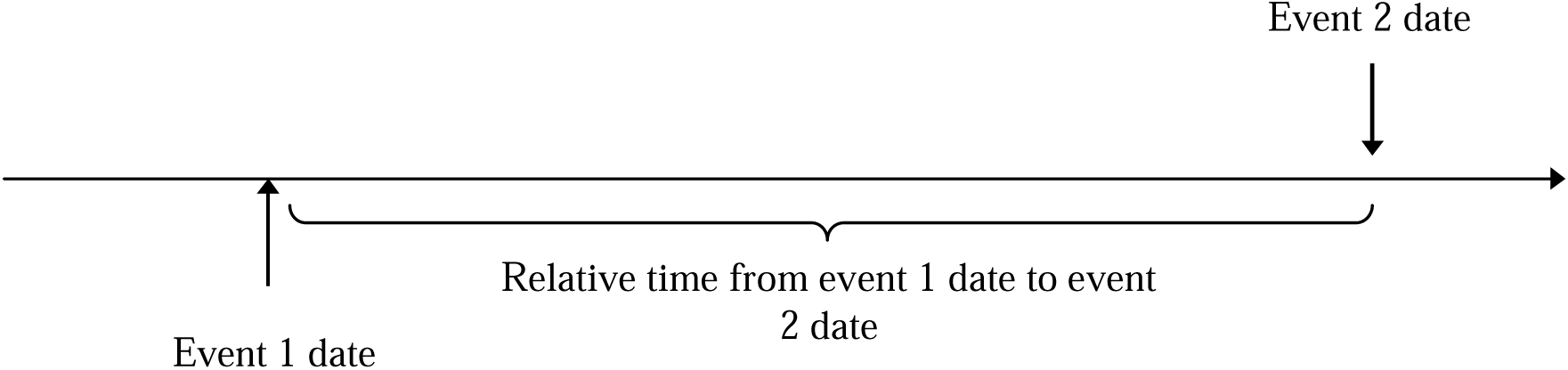

<u>Event 1</u>: first self-identification as sex worker; <u>Event 2</u>: first program contact

Among N=61 participants who ever been contacted by the program prior to interview date, we calculated the time from self-identification as sex worker to first program contact via direct measure based on Q805, and if there was missing; we then used indirect measure based on the time from first self-identification as sex worker to interview and the time from first program contact to interview to calculate the time between FSW to first program contact.

<u>Direct measure</u> (please see **Appendix Questions** for detailed questions and possible answers):

- Q805: Approximately how long after you first considered yourself a sex worker were you first contacted by peers/staff from an NGO, CBO OR FBO?

- N=38/61 reported a valid time since the participant first self-identified as sex worker to first program contact

<u>Indirect measure</u>

- Time from first self-identification as sex worker to interview and time from first program contact to interview (created in **A.2.2.1** and **A.2.2.2**)
  - N=19/23 participants’ time from self-identification as sex worker to first program contact were imputed

<u>Other questions</u> used to reduce missing values

- Q804: Approximately how long after you had your first paying client were you first contacted by peers/staff from an NGO, CBO OR FBO?
  - N=2/4 participants’ time from self-identification as sex worker to first program contact were imputed

**Final**: N = 59/61 participants who were <u>ever</u> contacted by a program reported a valid time from first self-identification as sex worker to program contact

### A.2.2.1 Time from first self-identification as sex worker to interview

<u>Event 1</u>: first self-identification as sex worker; <u>Event 2</u>: interview

<u>Main question</u> (please see **Appendix 1** for detailed questions and possible answers):

- B335: How many years (months) ago did you first consider yourself a sex worker? N=396 with non-missing values with N=12 missing

<u>Other questions used to reduce missing values</u>

- B336: How many years ago did you first enter sex work?
- B337: For how long have/had you been in sex work?
- Q318: How many years ago was it when you first had sex with a man where the price of sex was negotiated before the sex event?
- Q327: For how long have you been taking paying clients Following above order, N=9/12 missing was imputed

**Final**: N=405/408 participants have reported time from first self-identification as sex worker to interview N=3 missing

### A.2.2.2 Time from first program contact to interview

<u>Event 1</u>: first contacted by peers/staff from an NGO, CBO or FBO; <u>Event 2</u>: interview

<u>Question</u> (please see **Appendix 1** for detailed questions and possible answers):

- Q803: How many years (and months) ago were you first contacted by peers/staff from an NGO, CBO or FBO?

**Final**: N=56/61 participants who ever been contacted by a program by the time of interview have a valid time from first program contact to interview

### A.2.2.3 Time from self-identification of sex worker to <u>retirement of sex worker</u>

<u>Event 1</u>: self-identification of sex worker; <u>Event 2</u>: retirement of sex worker

<u>Question</u> (please see **Appendix 1** for detailed questions and possible answers):

- B333: How many years ago did you stop sex work?
- B335: How many years (months) ago did you first consider yourself a sex worker?

N=16 participants reported that they are retired sex workers

**Final**: N=16 participants reported a valid time from first self-identification as sex worker to retirement of sex work

## Appendix 3. Cohort time frame definition

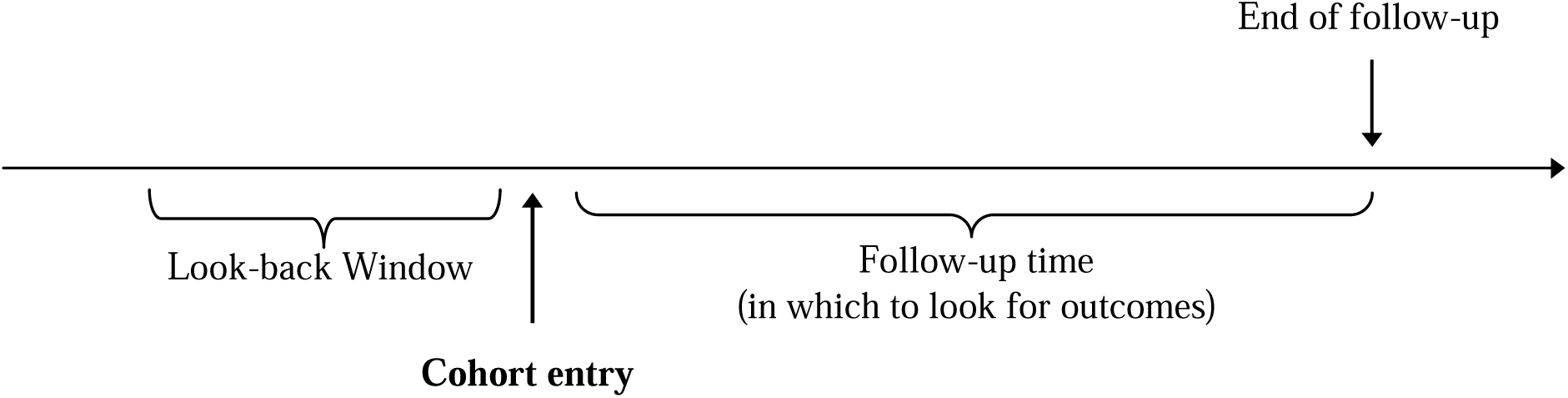

**Cohort entry**: self-identification of female sex worker

**End of follow-up**: <u>program contact</u>, <u>time of interview</u>, or <u>time of retirement from sex work</u> (whichever occurs the earliest)

**Step 1:** Participants who were <u>never</u> contacted by a program have end of follow-up of the <u>earlier</u> of:

- Interview date (N=332)
- Retirement of sex work (N=13)

**Step 2:** Participants were <u>ever</u> contacted by a program have end of follow-up of the <u>earlier</u> of:

- program contact (N=58)
- retirement of sex work (N=1)

**Step 3:** Participants could be contacted by program prior to index event (self-identification of female sex worker). N=12 participants were excluded in survival analyses since they were contacted by program during look-back window. These participants were included in estimating the prevalence of program contact among our study sample.

**Final**: N=4 participants have missing value for the follow-up time and N=392 participants reported a valid time from first self-identification as sex worker to retirement of sex work

## Appendix 4. Additional characteristics of study participants between those who have ever/never been contacted by program by time of study

| Characteristics (N(%<sup>a</sup>)) | Total (N = 408) | Contacted by program by time of study (N = 61) | Never contacted by program (N = 347) | P-value |
| --- | --- | --- | --- | --- |
| **CURRENT<sup>b</sup> PERCEPTIONS** |  |  |  |  |
| General perception of sex work<sup>i</sup> |  |  |  | 0.12 |
| Unpleasant (1-3) | 162 (39.7%) | 19 (31.1%) | 143 (41.2%) |  |
| Neutral (4-6) | 129 (31.6%) | 26 (42.6%) | 103 (29.7%) |  |
| Pleasant (7-10) | 111 (27.2%) | 15 (24.6%) | 96 (27.7%) |  |
| Self-perceived prestige of sex work<sup>ii</sup> |  |  |  | 0.32 |
| Not prestigious (1-3) | 225 (55.1%) | 29 (47.5%) | 196 (56.5%) |  |
| Neutral (4-6) | 94 (23%) | 15 (24.6%) | 79 (22.8%) |  |
| Prestigious (7-10) | 82 (20.1%) | 16 (26.2%) | 66 (19%) |  |
| Self-perceived danger of working in sex work<sup>iii</sup> |  |  |  | 0.39 |
| Dangerous (1-3) | 310 (76%) | 44 (72.1%) | 266 (76.7%) |  |
| Neutral (4-6) | 71 (17.4%) | 14 (23%) | 57 (16.4%) |  |
| Not dangerous (7-10) | 21 (5.1%) | 2 (3.3%) | 19 (5.5%) |  |
| Self-perceived lifestyle of working in sex work<sup>iv</sup> |  |  |  | 0.25 |
| Not desirable (1-3) | 189 (46.3%) | 26 (42.6%) | 163 (47%) |  |
| Neutral (4-6) | 67 (16.4%) | 7 (11.5%) | 60 (17.3%) |  |
| Desirable (7-10) | 144 (35.3%) | 27 (44.3%) | 117 (33.7%) |  |

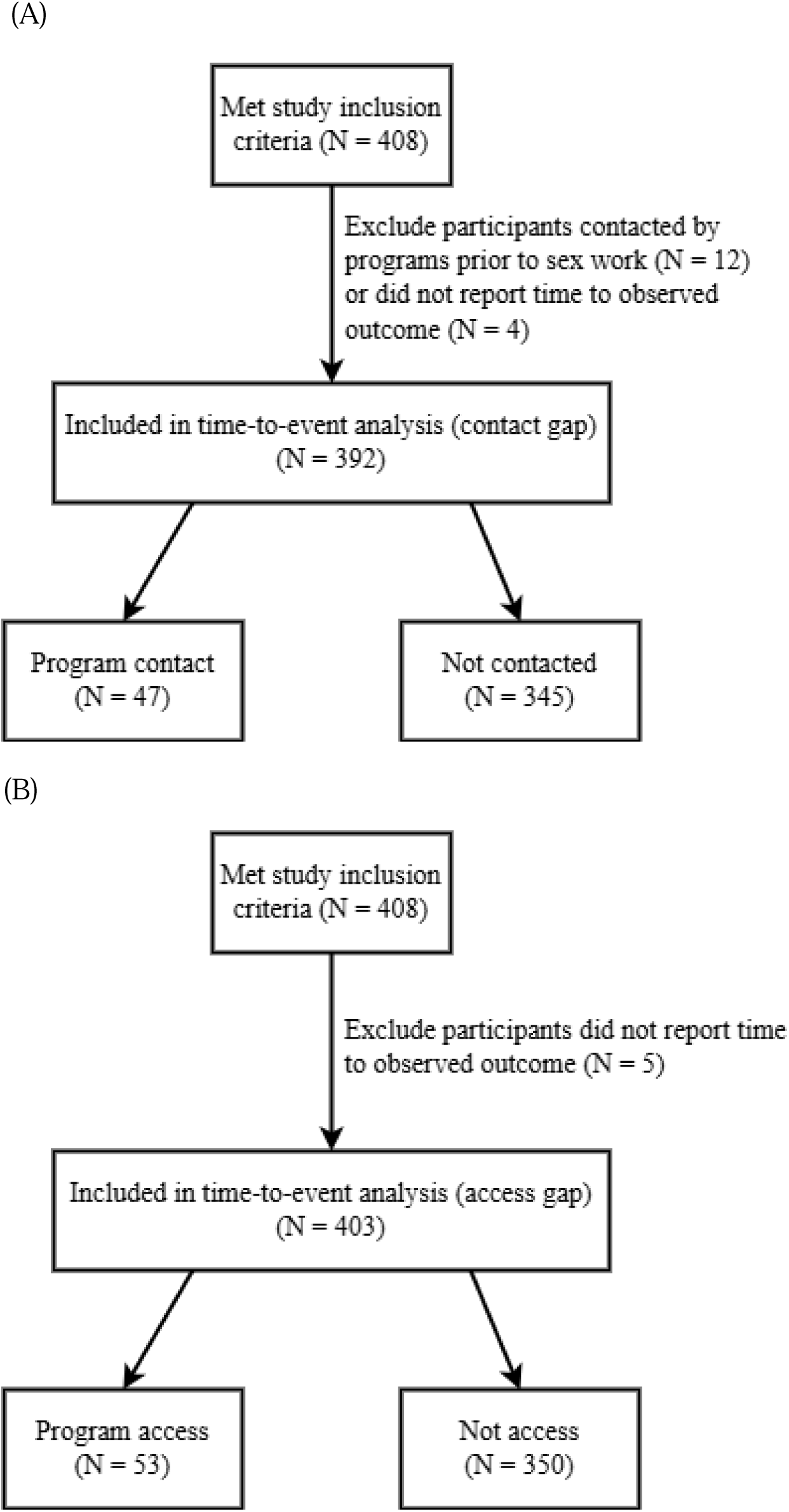

## Appendix 5. Flow charts of time-to-event analysis inclusion and exclusion criteria for (A) contact gap (primary outcome) and (B) access gap (secondary outcome)

(Sample size point estimate of YSW in Mombasa/Size of study sample included in time-to-event analysis and those contacted prior to sex work)*(Total person-months of follow-up of study sample/12), where the total person-months of follow-up was defined as time from self-identification as sex worker to program contact for those reported program contact, and time from self-identification as sex worker to survey date for those who did not report program contact.

## Appendix 6. Calculation of the minimum number of person-years of program contact delay among all young female sex workers in Mombasa

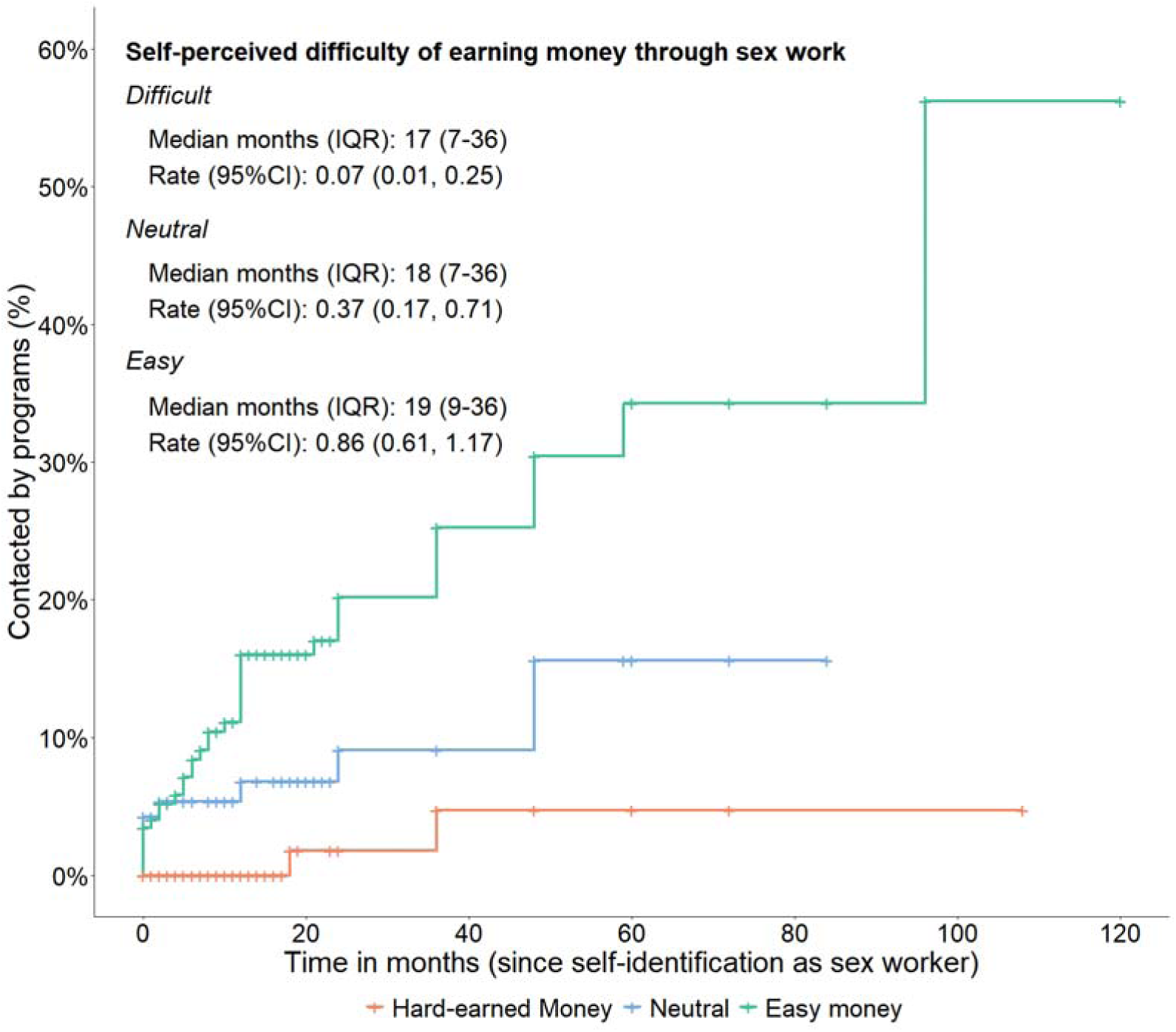

## Appendix 7. Kaplan-Meier curves of time from self-identification as sex worker to program contact by self-perceived difficulty of earning money through sex work. The graphs depict length of contact gap (time in months from initial self-identification as a sex worker to initial program contact). **Abbreviations:** CI: confidence interval; IQR: interquartile range

## Appendix 8. Characteristics of study participants between those who have ever/never accessed the program by time of study

| Characteristics (N(%<sup>a</sup>)) | Total (N = 408) | Accessed program (N = 56) | Never accessed program (N = 352) | P-value |
| --- | --- | --- | --- | --- |
| **CURRENT<sup>b</sup> PERCEPTIONS** |  |  |  |  |
| General perception of sex work<sup>i</sup> |  |  |  | 0.24 |
| Unpleasant (1-3) | 162 (40.3%) | 18 (32.7%) | 144 (41.5%) |  |
| Neutral (4-6) | 129 (32.1%) | 23 (41.8%) | 106 (30.5%) |  |
| Pleasant (7-10) | 111 (27.6%) | 14 (25.5%) | 97 (28%) |  |
| Self-perceived prestige of engaging in sex work<sup>ii</sup> |  |  |  | 0.28 |
| Not prestigious (1-3) | 225 (56.1%) | 26 (47.3%) | 199 (57.5%) |  |
| Neutral (4-6) | 94 (23.4%) | 14 (25.5%) | 80 (23.1%) |  |
| Prestigious (7-10) | 82 (20.4%) | 15 (27.3%) | 67 (19.4%) |  |
| Self-perceived danger of engaging in sex work<sup>iii</sup> |  |  |  | 0.68 |
| Dangerous (1-3) | 310 (77.1%) | 41 (74.5%) | 269 (77.5%) |  |
| Neutral (4-6) | 71 (17.7%) | 12 (21.8%) | 59 (17%) |  |
| Not dangerous (7-10) | 21 (5.2%) | 2 (3.6%) | 19 (5.5%) |  |
| Self-perceived lifestyle of engaging in sex work<sup>iv</sup> |  |  |  | 0.18 |
| Not desirable (1-3) | 189 (47.2%) | 22 (40%) | 167 (48.4%) |  |
| Neutral (4-6) | 67 (16.8%) | 7 (12.7%) | 60 (17.4%) |  |
| Desirable (7-10) | 144 (36%) | 26 (47.3%) | 118 (34.2%) |  |

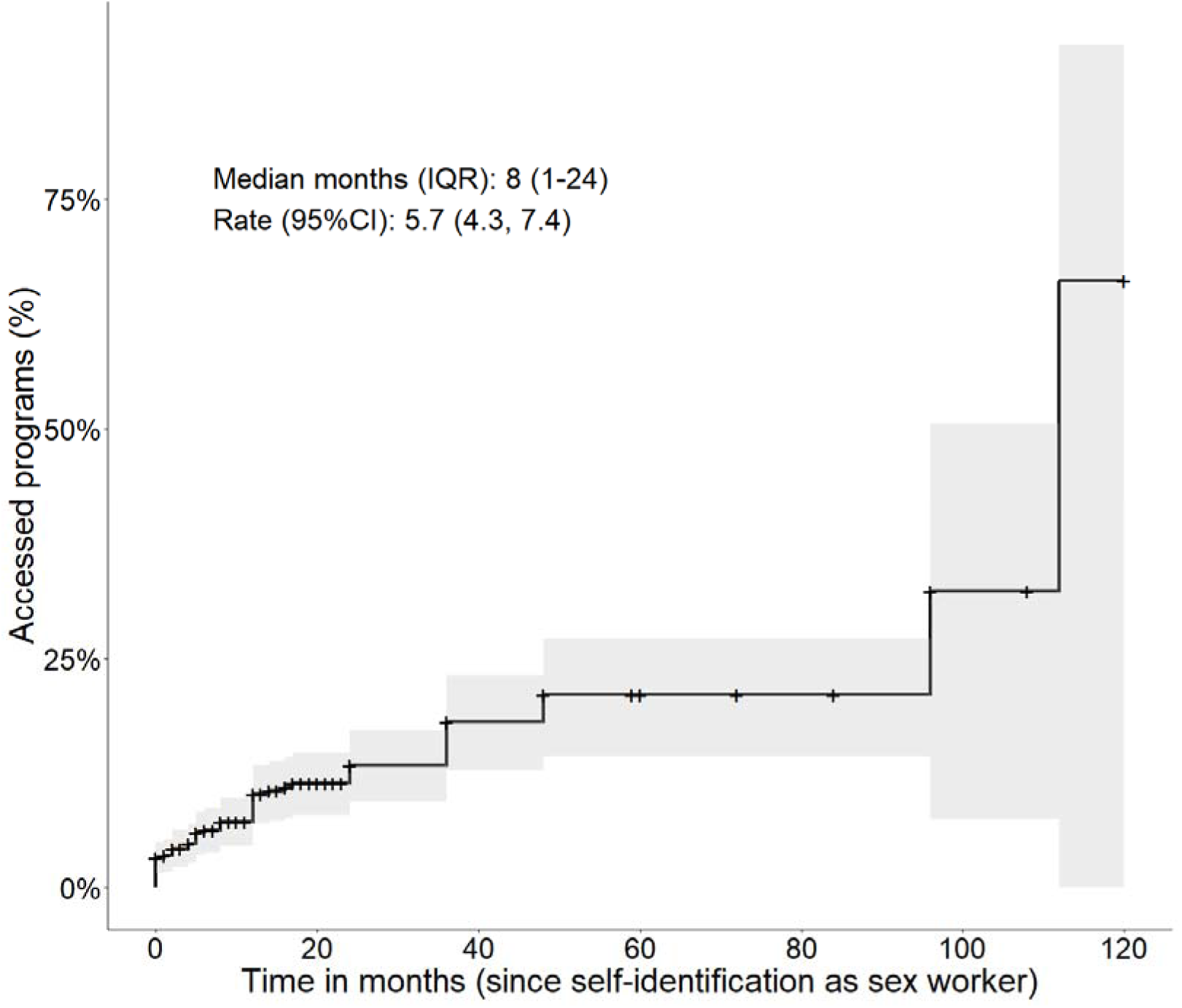

## Appendix 9. **Kaplan-Meier curve of time from self-identification as sex worker to program access for all participants.** The graph depicts length of access gap (time in months from initial self-identification as a sex worker to initial program access)

## Appendix 10. Non-adjusted and adjusted hazard ratios by determinants on access gap

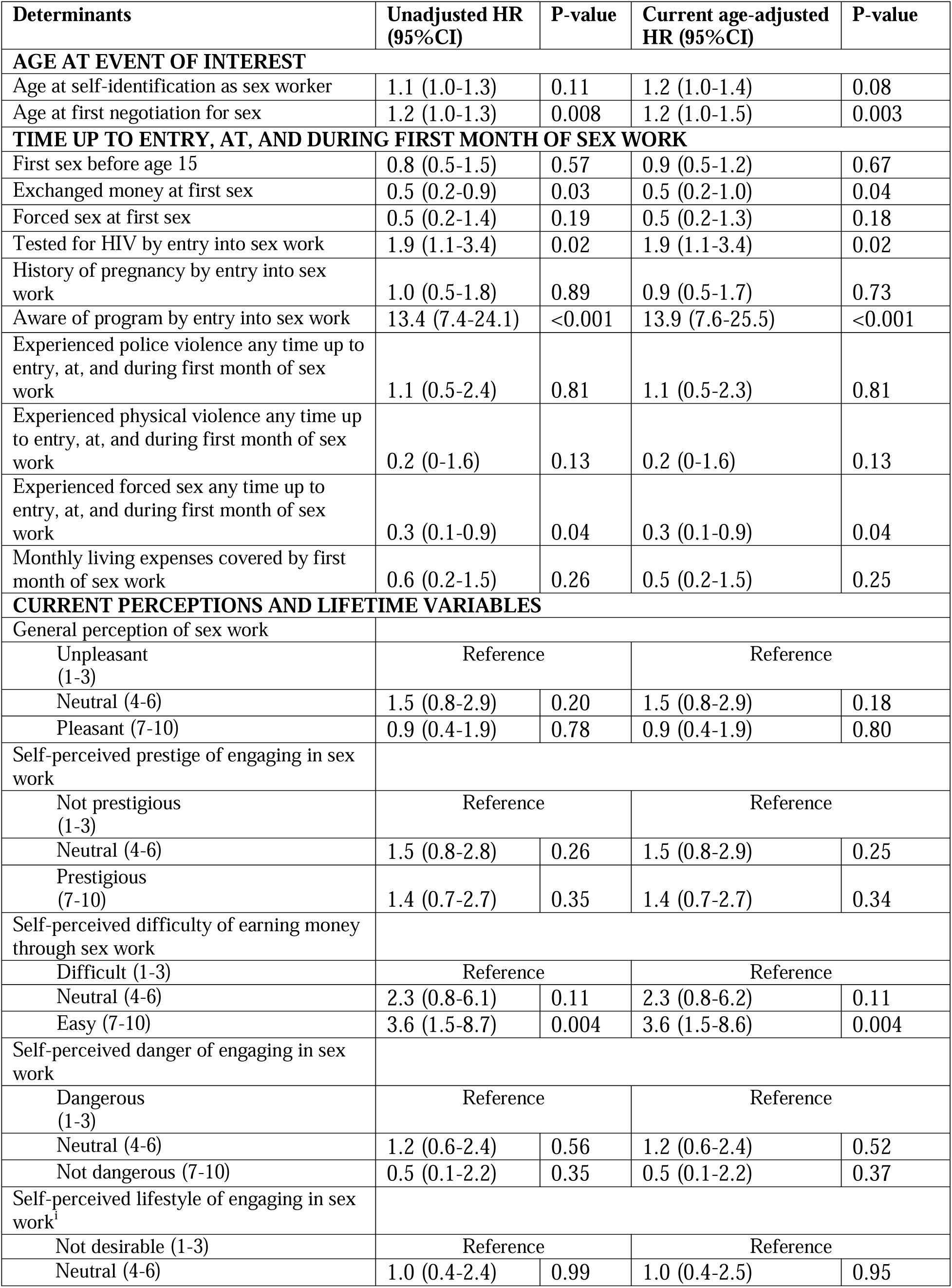
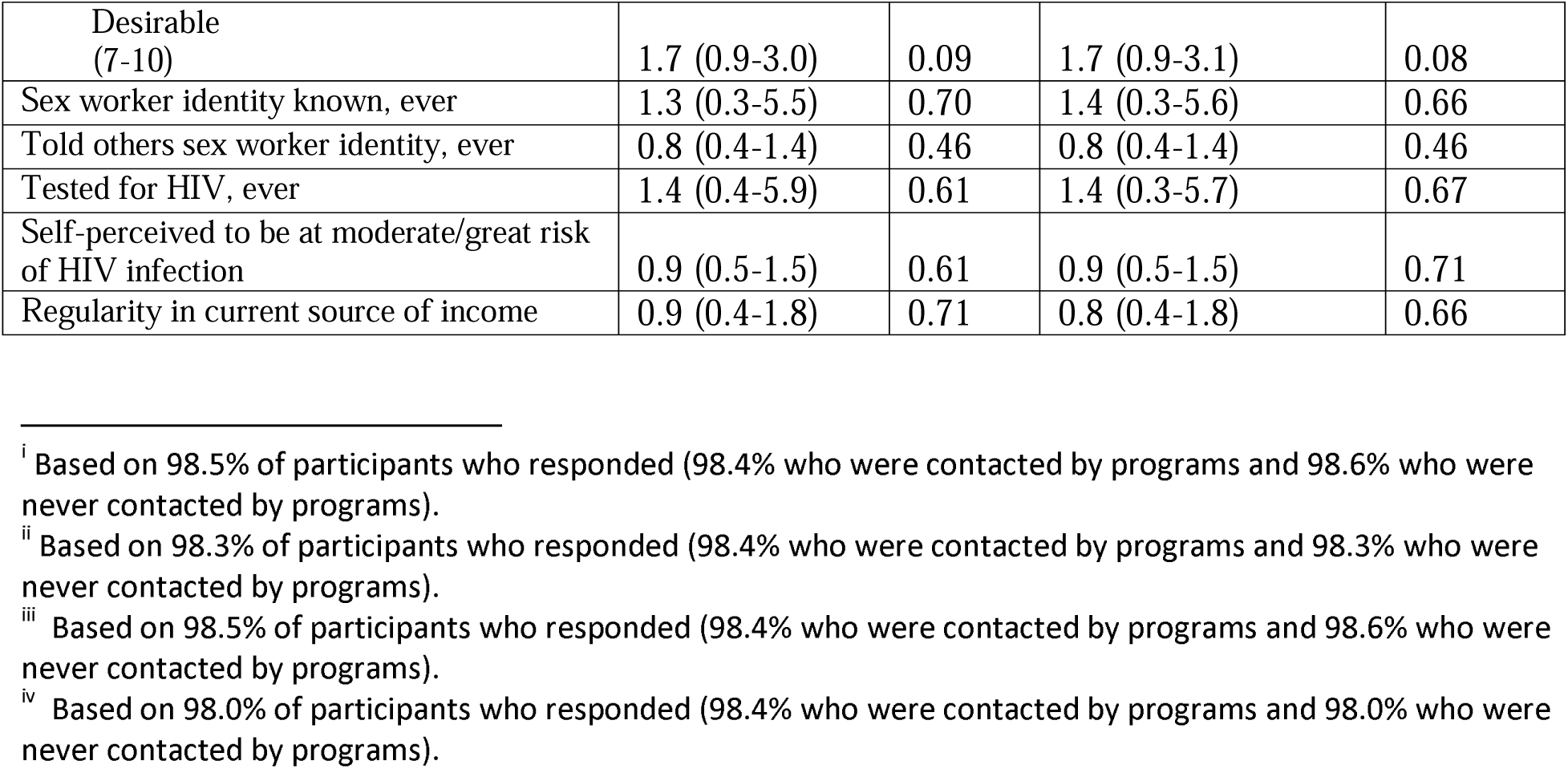

